# Sensitivity of endemic behaviour of Covid-19 under a multi-dose vaccination regime, to various biological parameters and control variables

**DOI:** 10.1101/2022.10.10.22280683

**Authors:** John Dagpunar, Chenchen Wu

## Abstract

For an infectious disease such as Covid-19, we present a new four-stage vaccination model (un-vaccinated, dose 1+2, booster, repeated boosters), which examines the impact of vaccination coverage, vaccination rate, generation interval, control reproduction number, vaccine efficacies, and rates of waning immunity, upon the dynamics of infection. We derive a single equation that allows computation of equilibrium prevalence and incidence of infection, given knowledge about these parameter and variable values. Based upon a 20 compartment model, we develop a numerical simulation of the associated differential equations. The model is not a forecasting or even predictive one, given the uncertainty about several biological parameter values. Rather, it is intended to aid qualitative understanding of how equilibrium levels of infection may be impacted upon, by the parameters of the system. We examine one at a time sensitivity analysis around a base case scenario. The key finding which should be of interest to policy makers, is that while factors such as improved vaccine efficacy, increased vaccination rates, lower waning rates, and more stringent non-pharmaceutical interventions might be thought to improve equilibrium levels of infection, this might only be done to good effect, if vaccination coverage on a recurrent basis, is sufficiently high.

## 1 Introduction

In this paper we introduce a mathematical model to explore the possible long-term prevalence of Covid-19 infection under a current 4-dose vaccination regime, which we refer to as doses 1 and 2, booster dose, and further booster doses. With varying vaccination coverage across the world, with newer variants of concern having much higher intrinsic transmissibility than the wild type, with immune escape and waning immunity both from vaccination and prior infection, and with different levels of non-pharmaceutical interventions (NPIs), this paper attempts to understand how these effect the long-term prevalence and incidence of infection. We cannot know the nature of future variants of concern, so this is not intended to be a predictive model. A central parameter is the control reproduction number (*R*_*c*_), that is the expected number of secondary infections from a primary infector in a completely susceptible population under specified NPIs.

We provide conditions for the disease to be eliminated, and conversely for it to exhibit the more likely endemic behaviour. At the time of writing, in most countries the omicron variant is dominant with very high intrinsic transmissibility. For example, Ito, Piantham, and Nishiura, 2022 estimated the effective reproduction number to be 3.19 (95% CI (2.82,3.61)) times that of the delta variant under similar epidemiological conditions. With the more transmissible sub-lineages BA4 and BA5, a basic reproduction number, *R*^(0)^ of at least 10 is likely. We assume a constant level for *R*_*c*_ below *R*^(0)^ to reflect a supposed new normal of ‘living with the virus’. We assume that this arises in such a way as to maximise expected utility to society. That might reflect balancing Covid mortality, economic damage, effects of a compromised health system, damage to young persons’ education, reduced individual freedom, etc. It assumes, for example, that during colder periods when contact between people is greater, that individual and community behaviour and any government interventions adjust themselves to maintain that constant level of *R*_*c*_.

A key feature of the paper is the derivation of a single equation which allows the easy computation of equilibrium prevalence of infection given estimates of the various parameter values. This aids understanding of how differing levels of vaccine coverage, immune escape both from vaccine and prior infection, and waning immunity, all impact upon the nature of endemic equilibrium. We produce specimen epidemic curves over five years. We show quantitatively how the cumulative infection incidence over five years varies with different parameter scenarios. We perform sensitivity analyses to show which biological parameters have the greatest impact upon equilibrium levels.

The structure of the paper is as follows. In section 2 we summarise the relevant literature in this area. In section 3, the mathematical model is developed. Experimental results for a base case scenario are given in section 4. In section 5 we conduct sensitivity analysis by perturbing the base case scenario. In section 6, we summarise and flag the importance of maintaining a high level of repeated boosters for a large proportion of the population.

## 2 Vaccine models reviewed

There is an established history of using mathematical models to study the evolution of epidemics and the effectiveness of vaccination programmes in epidemic control. In searching the literature, we were particularly interested in identifying those studies that included waning of immunity, which seems to be an important issue with Covid-19; see, for example, Altmann and Boyton, 2021.

Scherer and McLean, 2002 developed SIR vaccination models to answer basic questions regarding diseases such as measles, mumps, and rubella, where a certain proportion are vaccinated just once in early childhood, and where an assumption of lifelong immunity is relaxed by introducing waning of immunity. They show how to calculate thresholds for disease-free and endemic behaviour. If the threshold is only just exceeded, then that typically results in long honeymoon periods of low prevalence between successive subsequent waves. Feng, Towers, and Yang, 2011 develop SIR and stochastic simulation models for pandemic influenza with seasonal transmission rates, vaccination, and antiviral treatments included. As waning immunity is not modelled, the question of long-term endemic levels of disease is not considered.

When we focus on literature for Covid-19 and related vaccination programmes, SIR and SEIR models are frequently used. Hollingsworth, Okamoto, and Lloyd, 2020, in a non-immunising environment, examine the combined effect of transient controls and waning immunity, that can result in accumulation of susceptibles and a resultant emergence of larger than expected waves, the so-called divorce effect. Annas et al., 2020 and Batistela et al., 2021 both model vaccination by moving people from a susceptible compartment to a recovered one, which implicitly assumes 100% vaccine efficacy and both include theoretical stability analysis of disease-free and endemic equilibrium. Giordano et al., 2021 use a compartmental vaccine model to predict outcomes in Italy from initial roll-out at April 2021 until January 2022, under various Non-Pharmaceutical Intervention (NPI) control strategies and rates of vaccination of the population. Their results confirm the importance of not releasing NPIs until a sufficient proportion of the population has been vaccinated. The methodology optimistically assumes 100% vaccine efficacy and no waning of immunity after vaccination or infection.

Models developed by Xu, Wu, and Topcu, 2021, Ghostine et al., 2021, Wintachai and Prathom, 2021, and Antonini, Calandrini, and Bianconi, 2021 are age-homogeneous. In the early stages of the pandemic in the UK, Crellen et al., 2021 used an SEIR model to investigate the effect of different mean durations of immunity (90-365 days), on short-term dynamics and long-term endemic behaviour, in an age-based model, in the absence of vaccination, under various assumptions about the time-varying effective reproduction number. In Xu, Wu, and Topcu, 2021, the authors consider a short-term SEIR model over 200 days, that optimises vaccination roll-out, subject to thresholds on death rates, cumulative deaths, and target numbers of those vaccinated. It assumes 100% vaccine efficacy, with no waning of immunity following vaccination or infection. Ghostine et al., 2021 developed a SEIR model for short-term predictions in Saudi Arabia. It is age-homogeneous, fits to actual data, and takes account of vaccine efficacy, but not of waning immunity. Wintachai and Prathom, 2021 use a SEIR model and study the efficiency of vaccines for Covid-19 situations, again fitting parameters to actual data. Antonini, Calandrini, and Bianconi, 2021 use a mean duration of vaccinal and infection immunity of 240 days to predict short-term outcomes in Italy. For the first wave of Covid-19, Esteban and Almodovar-Abreu, 2021 use a 7 compartment vaccination model, which assumes no waning of efficacy, to forecast and compare deaths at 180 days into the first wave for assumed un-vaccinated and vaccinated populations under an assumed 95% vaccine efficacy. Steyn et al., 2022 develop a vaccination model for New Zealand, assuming no waning of efficacy, with an assumed basic reproduction number of 6 for the delta variant. They concluded that almost 100% vaccination coverage would be needed to allow controls to be removed, if the objective was to have subsequent occasional small outbreaks.

Other authors have used heterogeneous SIR/SEIR models. Saad-Roy et al., 2020 develop a Covid-19 SIR based model demonstrating a large range of model outcomes, ranging from elimination to high levels of endemic behaviour, as the speculative biological parameters, including vaccine efficacy and waning rates, are changed. The authors emphasise the importance of characterising these parameters if such models are to be useful for informing management policies. Moore et al., 2021 developed a compartmental model, heterogeneous by age and region, to predict the evolution of Covid-19 under various combinations of vaccine roll-out and time-dependent NPIs in the UK. This pre-dated the delta variant’s emergence as the dominant variant. It does not consider waning of immunity, either from infection or vaccination, or a combination of these two. We try to address these features, which are now recognised to be an important issue, as exemplified by a perceived urgent need to roll out booster doses. A finding of their study is that vaccination alone, even pre-delta, would not contain the virus. Patel et al., 2021 use agent-based simulation within an SEIR framework with heterogeneous transmission and age-specific mortality rates to model decision strategies over an 18-month period in North Carolina. The model does not consider waning immunity. The authors found that removing NPIs while distributing vaccines resulted in a significant increase in the number of infections, hospitalisations, and deaths.

Many of the above are characterised by being short-term predictive studies, prior to emergence of the delta and omicron variants, fitting parameters to local data, and with no explicit consideration of waning immunity and vaccine efficacy. There appears to be a dearth of modelling showing how people graduate from non-vaccinated status, to doses 1 and 2, through to boosters, and then to top-up boosters. This is a key feature of our model. It is now generally believed that Covid-19 will be with us for a long time, Scudellari, 2020. At the time of writing, estimates of the efficacy against infection by the delta variant were improving Pouwels et al., 2021, but there is uncertainty in waning rates and one cannot predict what new more transmissible variants might emerge. While writing the paper, the omicron variant became dominant and data on efficacy against infection, as opposed to symptoms, together with waning rates after doses 1 and 2, booster, and top-up boosters are not known with any great certainty. Therefore, rather than prediction, we are interested in understanding what the long-term endemic levels of infection might be under speculative assumptions about vaccine efficacy, waning immunity following both vaccination and infection, and control reproduction number. In our model we also include efficacy against transmission given infection, in the hope that future research may be able to elicit estimates of this.

## 3 A multi-dose vaccination model

The model is of a homogeneously mixing population with 20 compartments. Figure 1 shows the structure of the compartmental model. We assume that births and non-Covid-19 deaths can be ignored since the rates are small in comparison to other compartmental entering and leaving rates. There are four vertical sub-blocks, each comprising five compartments. These sub-blocks are identified as vaccination stages 0,1,2,3 respectively. Stage 0 contains persons who are currently non-vaccinated. Stage 1 contains those who have received only doses 1 and 2. For simplicity, doses 1 and 2 are grouped together, as from the start of the pandemic, in the UK at least, a person was not considered properly immunised until they had received both. Stage 2 contains persons vaccinated with doses 1,2, and a booster dose, but not a second booster. Stage 3 contains those who have received doses 1,2, booster and one or more subsequent boosters.

**Figure 1:**
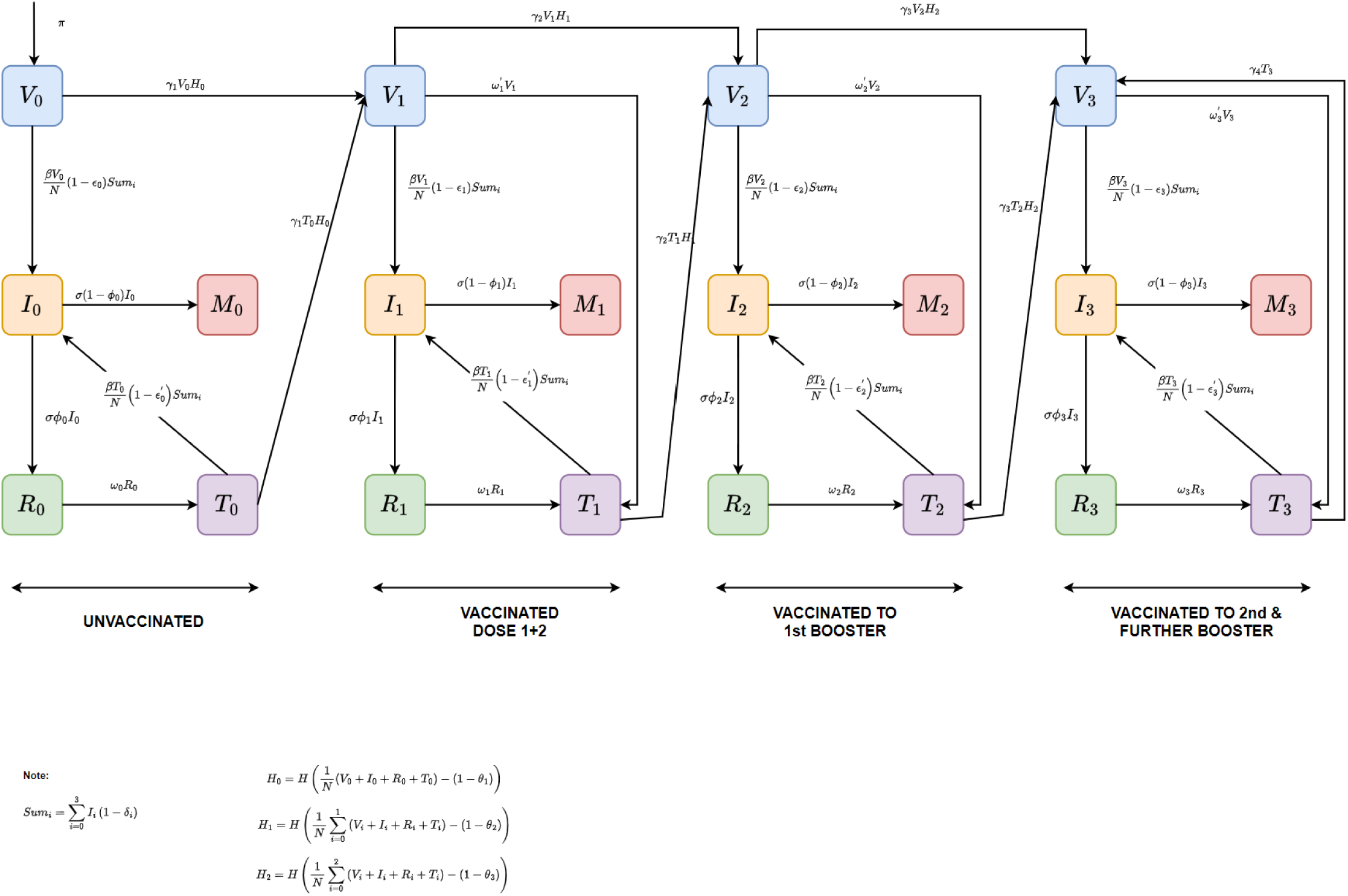
Flow chart of the vaccine model with variables and parameters shown in Table 1.

In vaccination stage 0, *V*_0_ is the current number of non-vaccinated persons who have not been infected. *I*_0_ is the prevalence of those infected. For simplicity, and to reduce the number of compartments, we assume that there is no latent period (exposed but not yet infectious) as we are concerned only with the long-term behaviour of the epidemic curve spanning many years. *R*_0_ is the number who have recovered from the most recent infection and considered to have full immunity initially. However, that immunity wanes at rate *w*_0_ so that *T*_0_ is the number who now do not have complete immunity but one that is equivalent to a vaccine with efficacy 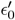. The probability of death given infection is 1 − *ϕ*_0_ and *M*_0_ is the cumulative number of deaths of the non-vaccinated. Those who are part of *V*_0_ or *T*_0_ can transition to vaccination with doses 1 and 2 at rate *γ*_1_, but only if that does not exceed the threshold proportion *θ*_1_ who aspire to doses 1 and 2. The model allows for repeat infections of an individual within stage, moving from compartments *I*_0_ to *R*_0_ to *T*_0_ to *I*_0_. At the time of writing, approximately 56 million of a UK population of 67 million had received doses 1 and 2, and so, as an example, we set *θ*_1_ = 56*/*67.

**Table 1:** Description of variables and parameters in the model where rates are per day.

| Variable /Parameter | Interpretation | Illustrative Values |
| --- | --- | --- |
| $N$ | Number of living population | $67 \times 10^6$ (UK) |
| $V_i$ | Number in vaccination compartment of stage $i$ ($i = 0, 1, 2, 3$) | $\mathbf{V}(\mathbf{0}) = (V_0(0), V_1(0), V_2(0), V_3(0)) = (N - 1, 0, 0, 0)$ |
| $I_i$ | Prevalence of infectious in vaccination stage $i$ ($i = 0, 1, 2, 3$). | $\mathbf{I}(\mathbf{0}) = (I_0(0), I_1(0), I_2(0), I_3(0)) = (1, 0, 0, 0)$ |
| $R_i$ | Number of recovered in vaccination stage $i$ ($i = 0, 1, 2, 3$). | $\mathbf{R}(\mathbf{0}) = (R_0(0), R_1(0), R_2(0), R_3(0)) = (0, 0, 0, 0)$ |
| $T_i$ | Number in vaccination stage $i$ with waned efficacy $\epsilon'_i$ ($i = 0, 1, 2, 3$). | $\mathbf{T}(\mathbf{0}) = (T_0(0), T_1(0), T_2(0), T_3(0)) = (0, 0, 0, 0)$ |
| $M_i$ | Cumulative number of Covid-19 deaths in vaccination stage $i$ ($i = 0, 1, 2, 3$). | $\mathbf{M}(\mathbf{0}) = (M_0(0), M_1(0), M_2(0), M_3(0)) = (0, 0, 0, 0)$ |
| $R_c$ | Control reproduction number=average number of secondary infections from one primary infector in a completely susceptible population, under current NPIs. | $R_c = 4$ |
| $\sigma$ | Rate of leaving the infected state. | $1/7$ |
| $\beta$ | Transmission rate for a completely susceptible population under current NPIs | $\beta = R_c \sigma$ |
| $\omega_i$ | Waning rate following infection. | $\omega_0 = \omega_1 = \omega_2 = \omega_3 = 1/180$ |
| $\omega'_i$ | Waning rate following vaccination. | $\omega'_1 = \omega'_2 = \omega'_3 = 1/360$ |
| $\epsilon_i$ | Peak efficacy against infection following vaccination ($i = 0, 1, 2, 3$). | $\epsilon_0 = 0, \epsilon_1 = 0.55, \epsilon_2 = 0.75, \epsilon_3 = 0.85$ |
| $\epsilon'_i$ | Waned efficacy against infection following compartment $T_i$ ($i = 0, 1, 2, 3$). | $\epsilon'_0 = 0.05, \epsilon'_1 = 0.25, \epsilon'_2 = 0.3, \epsilon'_3 = 0.4$ |
| $\delta_i$ | Efficacy against transmission given infection ($i = 0, 1, 2, 3$). | $\delta_0 = \delta_1 = \delta_2 = \delta_3 = 0$ |
| $1 - \phi_i$ | *Prob* (death given infection) ($i = 0, 1, 2, 3$). | $1 - \phi_0 = 2 \times 10^{-3}, 1 - \phi_1 = (1 - \phi_0) \times \frac{1-0.48}{1-0.15}, 1 - \phi_2 = (1 - \phi_0) \times \frac{1-0.9}{1-0.6}, 1 - \phi_3 = (1 - \phi_0) \times \frac{1-0.95}{1-0.7}$ |
| $\pi$ | Population replenishment rate | $\pi = \sigma \sum_{i=0}^3 (1 - \phi_i) I_i$ |
| $\gamma_i$ | Transition rate per person to next vaccination stage ($i = 1, 2, 3, 4$) | $\gamma_1 = \gamma_2 = \gamma_3 = \gamma_4 = 1/90$ |
| $\theta_1$ | Proportion of population who aspire to dose 1 and 2. | $56/67$ |
| $\theta_2$ | Proportion of population who aspire to first booster. | $42/67$ |
| $\theta_3$ | Proportion of population who aspire to the second and further boosters. | $6/67$ |

The dynamics for vaccination stages 1, 2, 3 are similar to those of stage 0, with minor differences. Efficacy after vaccination wanes at rate 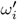 from *ϵ*_*i*_ to 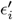. Following waning of a person’s immunity by either the vaccination or infection route, a person becomes a candidate for repeat infection or vaccination to the next stage. In stage 3 transition to the ‘next’ vaccination stage is actually within stage, to represent repeated booster doses. In all vaccination stages we allow for the possibility that the efficacy against transmission, *δ*_*i*_, improves with the number of vaccinations. By definition *δ*_0_ = 0 and we might expect *δ*_3_ ≥ *δ*_2_ ≥ *δ*_1_ ≥ *δ*_0_ = 0. Uncertainty about many parameter values, and indeed the current state of the system, leads us to emphasise that we do not intend this model to be predictive. Rather, we use it to gain some understanding of how different levels of aspiration to vaccination, of vaccine efficacy against infection and transmission, and of waning immunity, impact upon the long-term infection prevalence. By numerical simulation of the differential equations (1) we are also able to determine transient behaviour involving the various waves of the pandemic.

We cannot know the future course of the pandemic and the response in terms of new vaccines. For the moment, we assume the current situation of dose 1 plus 2, booster and subsequent boosters. Such subsequent boosters, possibly targeted to higher risk groups are assumed to continue in perpetuity. Indeed, in the UK third boosters are now being administered to such groups. We cannot predict the nature of future variants and while two or more may co-exist for a certain period, the more transmissible one is likely to dominate quickly, as happened with omicron superseding delta and delta dominating alpha. In future, with a different circulating variant, the model can be run with new parameter values in so far as they are known. Parameter definitions are shown in the second column of table 1. The epidemic dynamics shown in figure 1 lead to the differential equations (1).

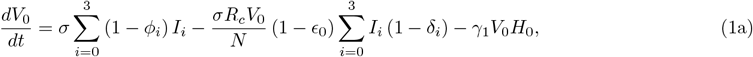

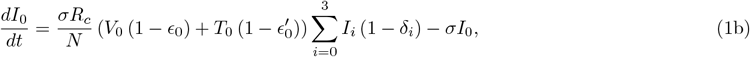

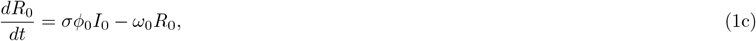

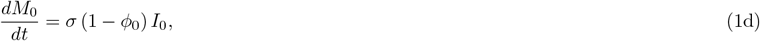

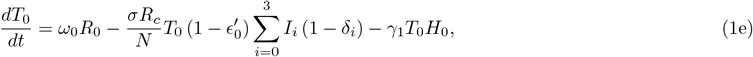

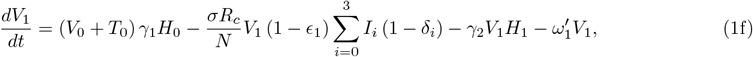

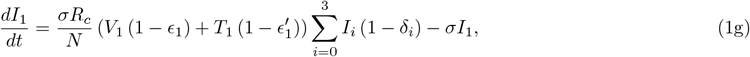

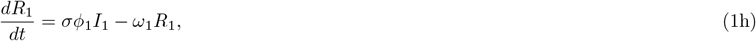

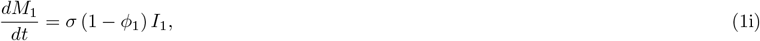

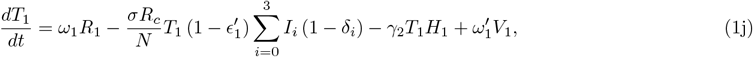

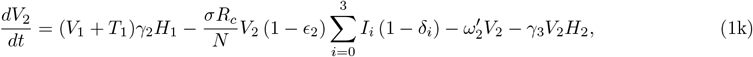

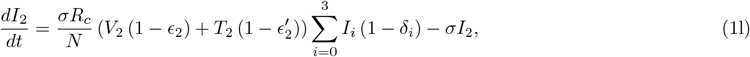

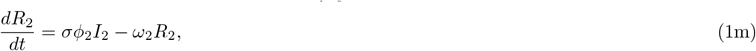

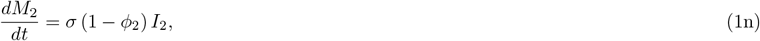

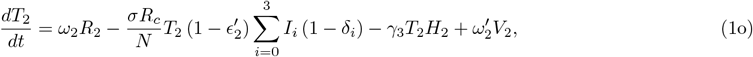

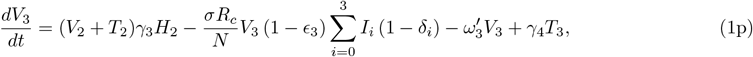

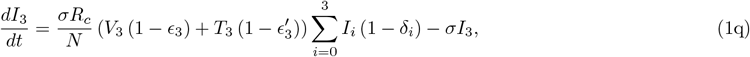

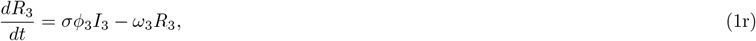

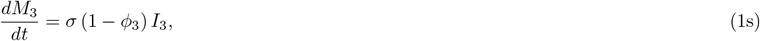

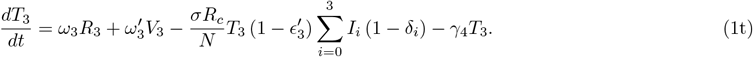

In the model differential equations (1),

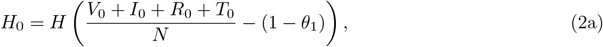

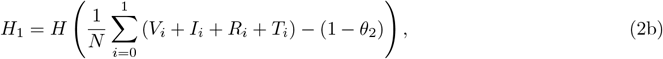

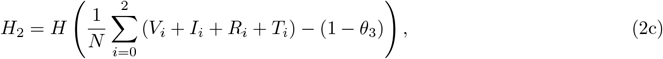

where the Heavyside function *H* is replaced by the continuous error function with very small scale parameter, that is

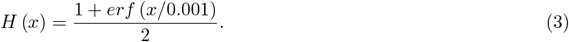

The control reproduction number *R*_*c*_ is the average number of secondary infections from a primary infector, under current non-pharmaceutical interventions (NPIs), in a hypothetical completely susceptible population. It is smaller than the basic reproduction number *R*^(0)^ because of the NPIs. In seeking to understand long-term dynamics we assume that *R*_*c*_ does not vary with time. That is, for *t >* 0 the mix of NPIs at any time, including individual behaviours such as level of homeworking, mask wearing (see for example Chu et al., 2020), improved ventilation, improved test-trace-isolate methods, adjust themselves in such a way that results in a constant *R*_*c*_, that is smaller than *R*^(0)^. Thus, during winter periods with more indoor mixing, the intrinsic transmission rate and therefore *R*^(0)^ might be expected to increase, but we assume a behavioural control policy that increases NPIs to bring *R*_*c*_ back to a constant level. The basic reproduction number for the omicron variant is perhaps as high as 12 but we will examine the impact of values of *R*_*c*_ below that, to reflect the new normal of ‘living with the virus’. We assume that there will be no new variant that is more transmissible, more virulent, has more vaccine escape than the omicron variant. In future as such changes become apparent, the parameter values can be reset. We do not intend the model to be predictive, rather to use it to gain some understanding of the impact of different levels of vaccination coverage, of waning immunity, and of vaccine efficacy, on the long-term infection prevalence and incidence. For these reasons, parameter values are to be regarded as illustrative ones only.

The size of the living population is an assumed constant 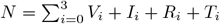 To achieve this it is assumed that the population replenishment rate 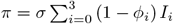. The proportion of living population in each compartment is scale invariant with respect to *N*. Henceforth, unless otherwise stated, we set *N* = 1 with *V*_*i*_, *I*_*i*_, *R*_*i*_, *T*_*i*_, *M*_*i*_ for *i* = 0, 1, 2, 3 now representing proportions of that living population. For this compartmental model the control reproduction number is

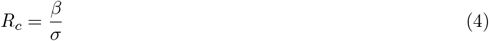

The expected transmission rate for a randomly selected infectious person is 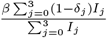 and this leads to an effective (time-dependent) reproduction number of

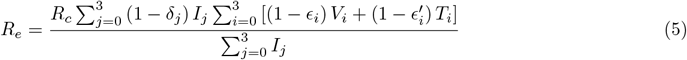

Let 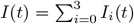 denote the total infection prevalence at time *t*. Then using the differential equations and (5) leads to

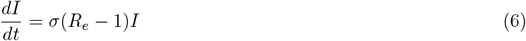

showing that the prevalence of infectives is increasing or decreasing according as *R*_*e*_ *>* 1, *R*_*e*_ *<* 1. Further, the cumulative incidence of infection in [0, *T*] is

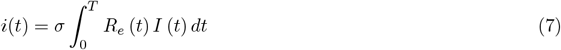

A disease free equilibrium (DFE) is obtained by setting the derivatives of (1a)-(1t) to zero with *I*_*i*_ = 0 for *i* = 0, 1, 2, 3. We find that the solution is

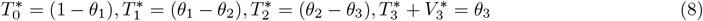

where 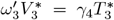 and all other state variables become zero. This corresponds to exactly meeting the vaccination aspirations, with static behaviour in all but vaccination stage 3 where further boosters are given once waning has set in.

In the absence of studies under current vaccine technology giving conclusive evidence that the efficacy against transmission given infection is anything other than zero, we consider a default scenario *δ*_3_ = *δ*_2_ = *δ*_1_ = *δ*_0_ = 0. In that case

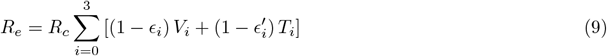

and from result (8), the disease free equilibrium is achieved when

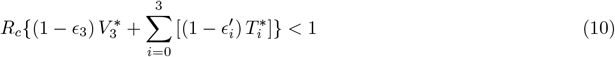

This is is the condition for disease elimination in the long-run. If it is not satisfied then the disease is endemic and persists for ever.

The more likely endemic equilibrium can be obtained by numerical simulation of the differential equations. However, when efficacy against transmission given infection is zero, we now derive a single equation which allows us to obtain the equilibrium prevalence of infection *I*, under the additional assumption that the population replenishment rate 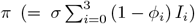 can be neglected. As we shall see in section 5, there is little difference between the numerically simulated equilibrium prevalence using the infection fatality rates shown in table 1 and the theoretical result assuming zero infection fatality rates, using equation 14 below. Defining *θ*_0_ = 1 and setting the derivatives to zero, we find that at equilibrium

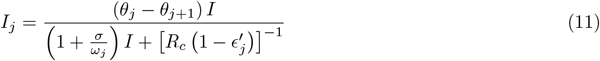

for *j* = 0, 1, 2 and

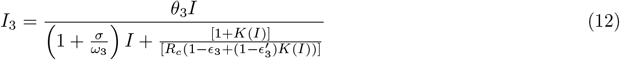

where

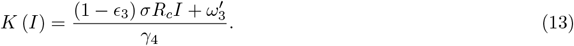

Adding these equations and equating to *I* we have

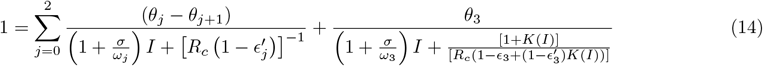

giving the equilibrium infection prevalence *I* in the endemic case. The threshold for this behaviour is obtained by setting *I* = 0 in (14), that is

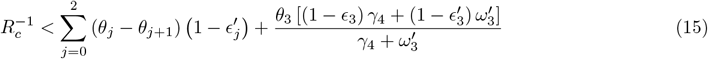

which is the complement of (10).

## 4 Experimental results for a base case scenario

We define a base case scenario using parameter values shown in column 3 of table 1. Those for vaccine efficacy are partly guided by the UK Health Security Agency, 2022 report, but as with other parameter values should be taken as a fictitious scenario, in view of uncertainties, the fact that the report refers to symptomatic infection (rather than symptomatic plus asymptomatic infection), the varying data for different combinations of vaccines, and no knowledge as to the nature of future variants and vaccines. The base case value for *σ* reflects a study by Manica et al., 2022 who estimated the mean generation interval for omicron as 6.84 days (95% CI 5.72-8.60). The base case figures for vaccine aspiration are guided by data released by the Public Health England, 2022 website as of 3 August 2022. In this simulation, all state variables are initially zero, except for *V*_0_(0) = (*N* − 1) */N* and *I*_0_(0) = 1*/N* where *N* = 6.7 *×* 10^7^ for the UK. We solve the system of differential equations (1) numerically for *t* = 365 *×* 5 days to illustrate the long-term behaviour of the state variables {*V*_*i*_, *I*_*i*_, *R*_*i*_, *T*_*i*_}. We have not attempted to show the dynamics of 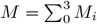. This would require accurate estimates of {1 − *ϕ*_*i*_} which will be dependent upon the age distribution within each stage and we might expect this to change with time as vaccines are rolled out. We concentrate on the behaviour of {*V*_*i*_, *I*_*i*_, *R*_*i*_, *T*_*i*_} and found it to be largely unaffected by changes in {1 − *ϕ*_*i*_} over a range of plausible values.

Figure 2 shows the dynamics for vaccination stages 0, 1, 2, and 3. We observe that the proportion of the population who are un-vaccinated (stage 0), decreases and reaches the equilibrium value of 16.2%, which is close to 1 −*θ*_1_ = 1 − 56*/*67 = 16.4%. Correspondingly, the proportion in stage 1 increases initially, and then decreases to an equilibrium *θ*1 − *θ*2 = 14*/*67 = 20.9% due to their migration to stage 2 (first booster). After around 500 days the proportion in stage 2 (first booster only) has stabilised at *θ*_2_ − *θ*_3_ = 36*/*67 = 53.7%, leaving 9.2% in stage 3 equilibrium.

**Figure 2:**
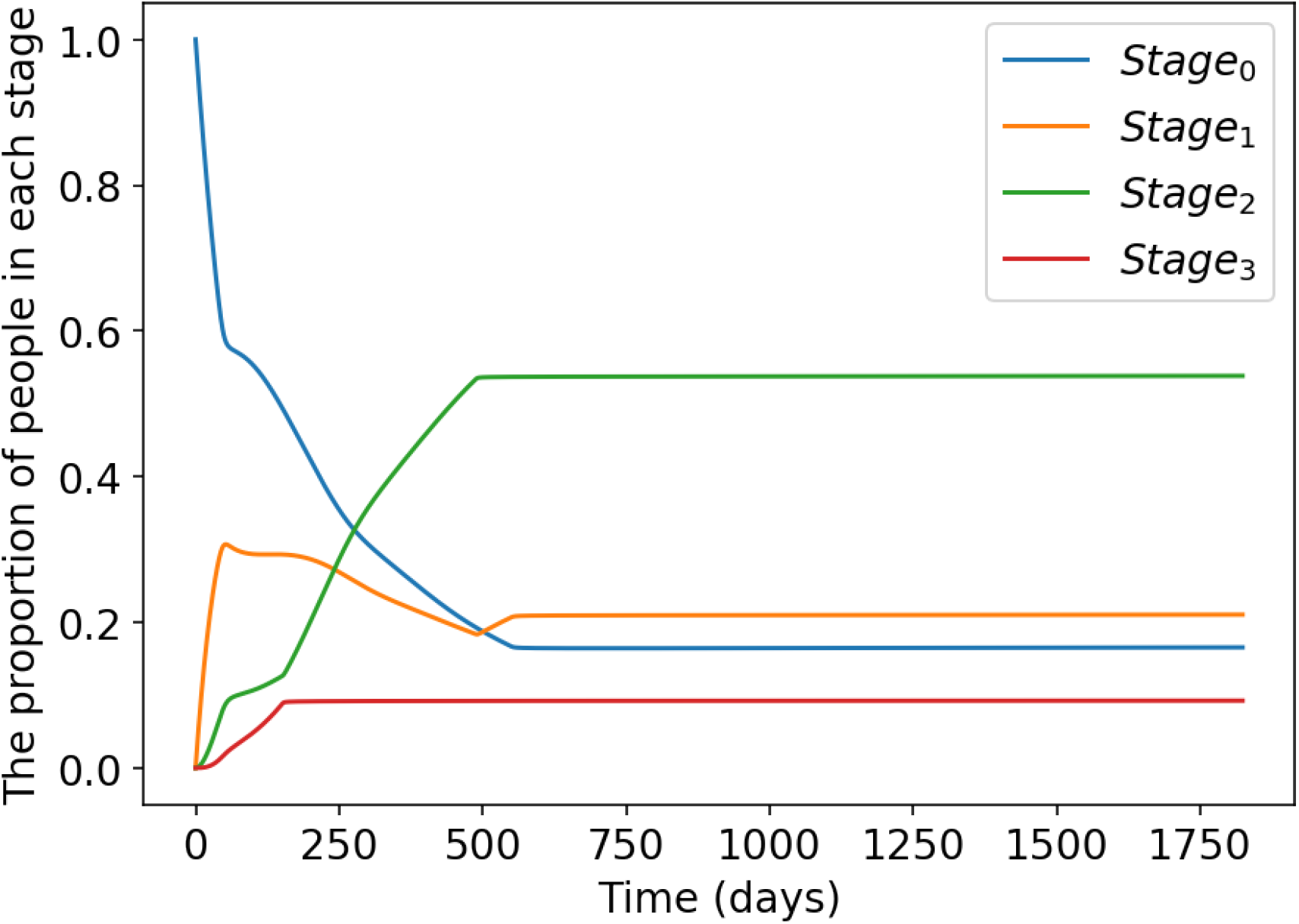
The proportion of people in each stage over time.

Figure 3 shows the changing proportion of people in each of the four categories of compartments. Thus, *V* (*t*) = *V*_0_(*t*) + *V*_1_(*t*) + *V*_2_(*t*) + *V*_3_(*t*), and similarly for the remaining 3 categories. As shown in figure 3, *V* (*t*) declines from an initial 100% (initially everyone is susceptible). It first decreases very sharply due to a huge first wave of infection followed by a short period of increase, due mainly to dose 1 +2 vaccination. It then declines gradually to an equilibrium level.The behaviour of the recovered fraction, *R*(*t*), is almost exactly opposite to that of *V* (*t*). The waned fraction *T* (*t*) increases following the first wave of infection, then decreases slightly, only to increase following the second wave, towards its equilibrium level. At equilibrium the prevalence of infection is approximately 2.4% of the population.

**Figure 3:**
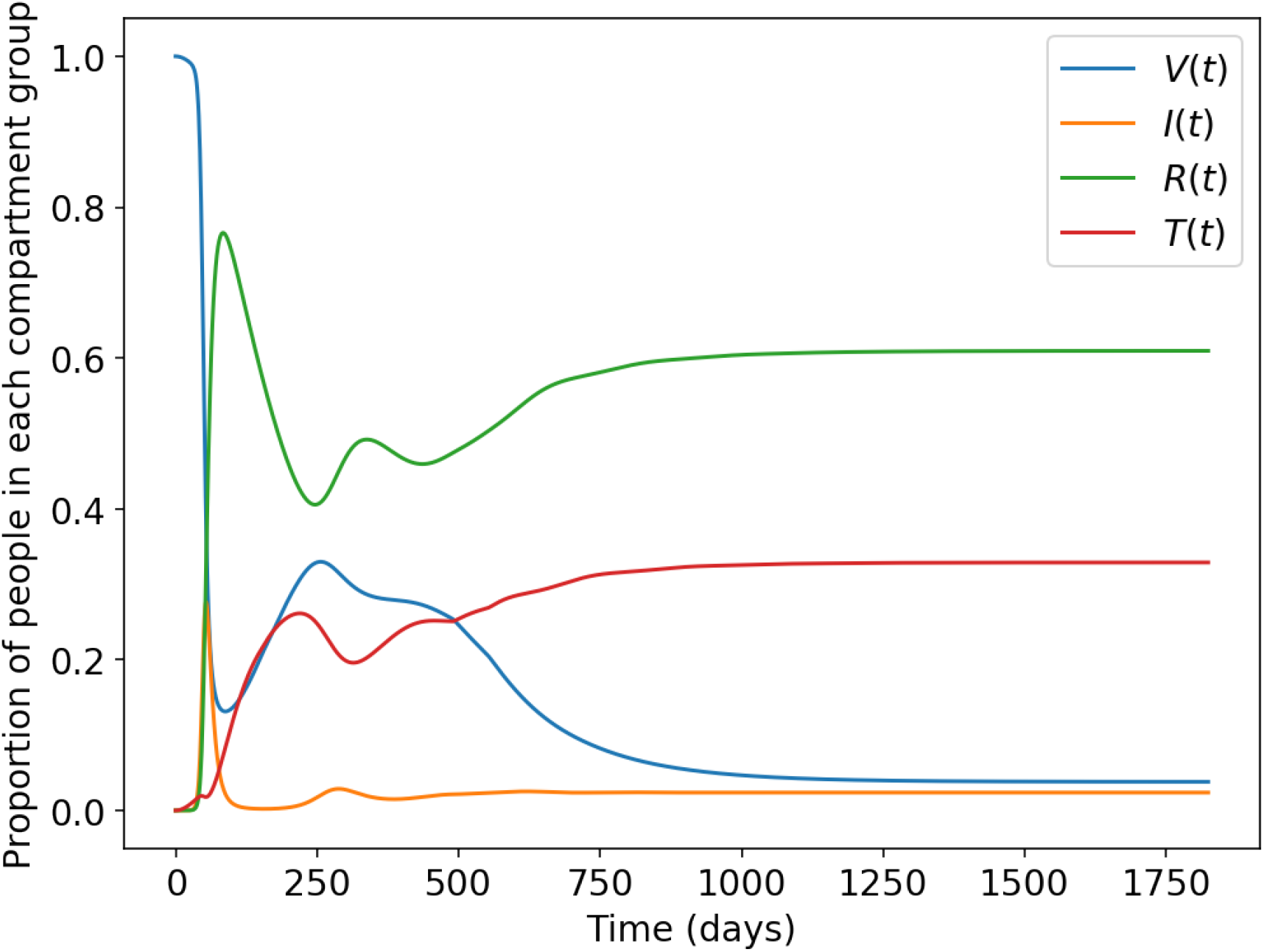
The proportion of people in each category of compartment over time.

Figures 4(a) and 4(b) show how the prevalence of infection, *I*(*t*), and its component parts *I*_0_(*t*), *I*_1_(*t*), *I*_2_(*t*), *I*_3_(*t*) change with time. From 4(a), a huge first wave dominated by *I*_0_(*t*) and to a lesser extent by *I*_1_(*t*) peaks at around 55 days. The second wave for all four components of infection is smaller than the first. As the prevalences, *I*_2_(*t*) and *I*_3_(*t*) are much smaller than the other two, figure 4(b) shows zoomed images of these two. In all cases, the honeymoon period between first and second peaks is larger than the time to first peak.

**Figure 4:**
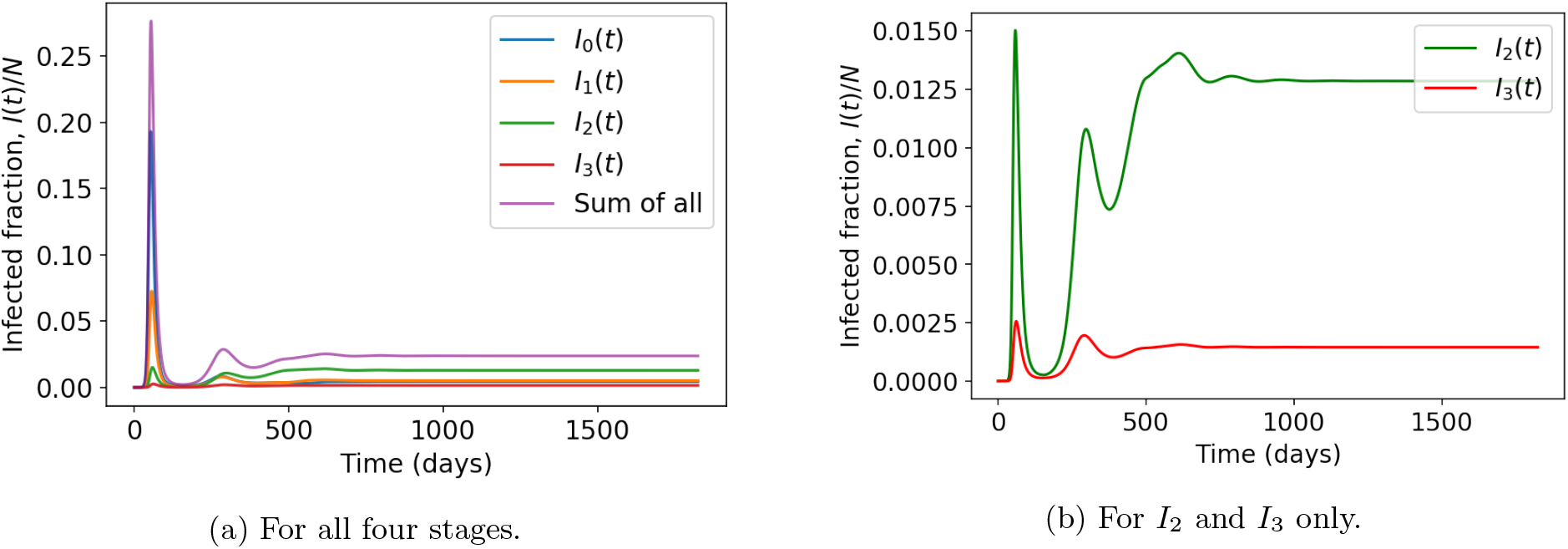
The change of infected fraction over time

Figure 5 shows the change of *R*_*e*_(*t*). *R*_*e*_ decreases sharply for about 70 days and reaches to its lowest value which is around 0.47. After that, it increases for around 200 days and then fluctuates around 1. After day 500 it remains very close to 1, reflecting equilibrium prevalence of infection.

**Figure 5:**
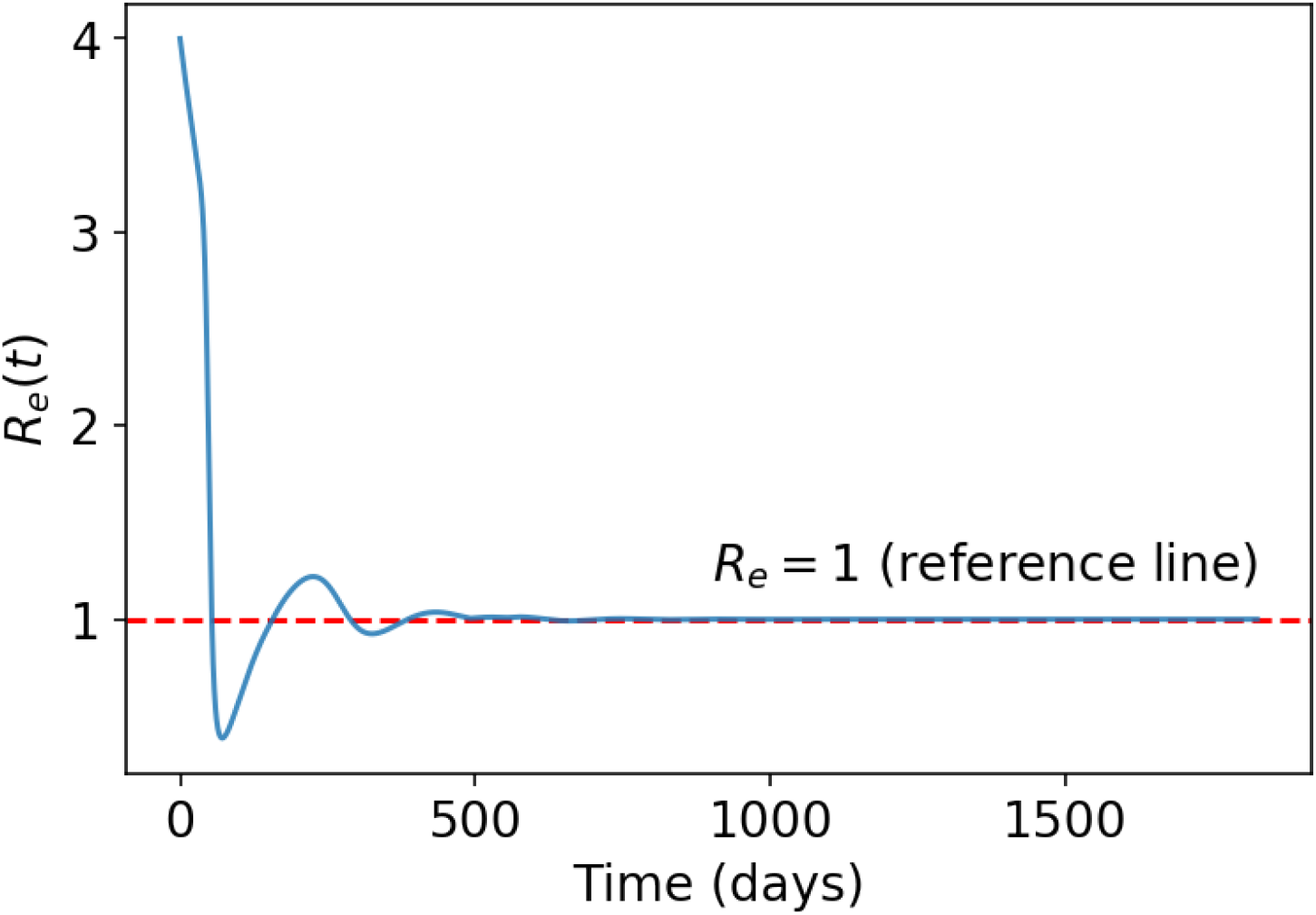
The change of *R*_*e*_(*t*).

## 5 Sensitivity analysis

The model requires input values for parameters. In general, estimates of biological parameters have high uncertainty. Here we perform sensitivity analysis for a range of scenarios, by one at a time change of parameter values from the base case scenario of section 4.

Table 2 shows corresponding equilibrium prevalence levels of infection, equilibrium incidence per year, and cumulative infections in (0,1825) days, the latter being a measure of the total damage inflicted by the virus over that period. In table 2, the “Theo.” column gives the theoretical equilibrium prevalence obtained from equation (14), while the “Exp.” column gives the experimental prevalence at *t* = 1825 in the numerical simulation of the differential equations. These are in close agreement. Apart from the changing values shown in the first column, all parameter values are those defined in the base case scenario (excluding *θ*) of table 1. The results shown in columns 2 to 5 are for 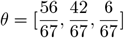 while those in columns 6 to 9, are those for a highly vaccinated population with *θ*= [1, 0.95, 0.9]. Figures 6 to 11 show the dynamic behaviour of infected prevalence while changing values of the parameter(s) noted in each caption. Those sub-plots labelled “(1)” are for 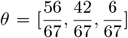 and those with label “(2)” are for *θ* = [1, 0.95, 0.9].

**Table 2:** A summary of equilibrium infection prevalence and incidence and cumulative infections over 5 years, under different conditions. The basic parameters are shown in Table 1

| Parameter<br>value | (1) $\theta = [\frac{56}{67}, \frac{42}{67}, \frac{6}{67}]$ | | | | (2) $\theta = [1, 0.95, 0.9]$ | | | |
| --- | --- | --- | --- | --- | --- | --- | --- | --- |
|  | Equilibrium<br>prevalence (%) |  | Equilibrium<br>incidence<br>per year<br>(/N) | Cumulative<br>infections<br>(/N) | Equilibrium<br>prevalence (%) |  | Equilibrium<br>incidence<br>per year<br>(/N) | Cumulative<br>infections<br>(/N) |
|  | Theo. | Exp. |  |  | Theo. | Exp. |  |  |
| a) Change $\sigma^{-1}$ | | | | | | | | |
| 7 | 2.37 | 2.37 | 1.24 | 6.30 | 0.52 | 0.49 | 0.27 | 2.15 |
| 14 | 4.57 | 4.57 | 1.19 | 5.95 | 1.00 | 0.96 | 0.26 | 1.68 |
| 3.5 | 1.21 | 1.21 | 1.26 | 6.48 | 0.27 | 0.25 | 0.29 | 2.37 |
| b) Change $\omega^{-1}$ and $\omega'^{-1}$ | | | | | | | | |
| $\omega^{-1} = 180, \omega'^{-1} = 360$ | 2.38 | 2.37 | 1.24 | 6.30 | 0.52 | 0.49 | 0.27 | 2.15 |
| $\omega^{-1} = 180, \omega'^{-1} = 180$ | 2.38 | 2.39 | 1.24 | 6.48 | 1.04 | 1.04 | 0.54 | 3.48 |
| $\omega^{-1} = 90, \omega'^{-1} = 360$ | 4.61 | 4.60 | 2.40 | 11.90 | 1.26 | 1.22 | 0.66 | 5.01 |
| $\omega^{-1} = 90, \omega'^{-1} = 180$ | 4.63 | 4.63 | 2.41 | 12.10 | 2.22 | 2.20 | 1.16 | 7.11 |
| $\omega^{-1} = 45, \omega'^{-1} = 360$ | 8.65 | 8.65 | 4.49 | 22.17 | 3.43 | 3.38 | 1.79 | 12.00 |
| $\omega^{-1} = 45, \omega'^{-1} = 180$ | 8.69 | 8.68 | 4.53 | 22.36 | 4.76 | 4.75 | 2.48 | 14.63 |
| c) Change $\epsilon = [\epsilon_0, \epsilon_1, \epsilon_2, \epsilon_3]$ | | | | | | | | |
| $[0, 0.55, 0.75, 0.85]$ | 2.37 | 2.37 | 1.24 | 6.30 | 0.52 | 0.49 | 0.27 | 2.15 |
| $[0, 0.3, 0.4, 0.7]$ | 2.42 | 2.42 | 1.26 | 6.82 | 1.53 | 1.54 | 0.80 | 5.00 |
| $[0, 0.3, 0.4, 1]$ | 2.28 | 2.27 | 1.18 | 6.39 | 0 | 0 | 0 | 1.61 |
| $[0, 1, 1, 1]$ | 2.28 | 2.25 | 1.18 | 4.81 | 0 | 0 | 0 | 0.50 |
| d) Change $\epsilon' = [\epsilon'_0, \epsilon'_1, \epsilon'_2, \epsilon'_3]$ | | | | | | | | |
| $[0.05, 0.25, 0.3, 0.4]$ | 2.37 | 2.37 | 1.24 | 6.30 | 0.52 | 0.49 | 0.27 | 2.15 |
| $[0.1, 0.3, 0.4, 0.5]$ | 2.21 | 2.21 | 1.15 | 5.88 | 0.18 | 0.04 | 0.10 | 1.41 |
| e) Change $\gamma^{-1}$. | | | | | | | | |
| 90 | 2.37 | 2.37 | 1.24 | 6.30 | 0.52 | 0.49 | 0.27 | 2.15 |
| 180 | 2.40 | 2.40 | 1.25 | 6.46 | 1.16 | 1.11 | 0.60 | 4.85 |
| 45 | 2.35 | 2.34 | 1.22 | 6.26 | 0.03 | 0 | 0.02 | 0.70 |
| f) Change $R_c$ | | | | | | | | |
| 4 | 2.37 | 2.37 | 1.24 | 6.30 | 0.52 | 0.49 | 0.26 | 2.15 |
| 6 | 2.83 | 2.83 | 1.48 | 7.79 | 1.78 | 1.63 | 0.93 | 5.86 |
| 8 | 3.06 | 3.06 | 1.60 | 8.55 | 2.39 | 2.40 | 1.25 | 7.63 |

**Figure 6:**
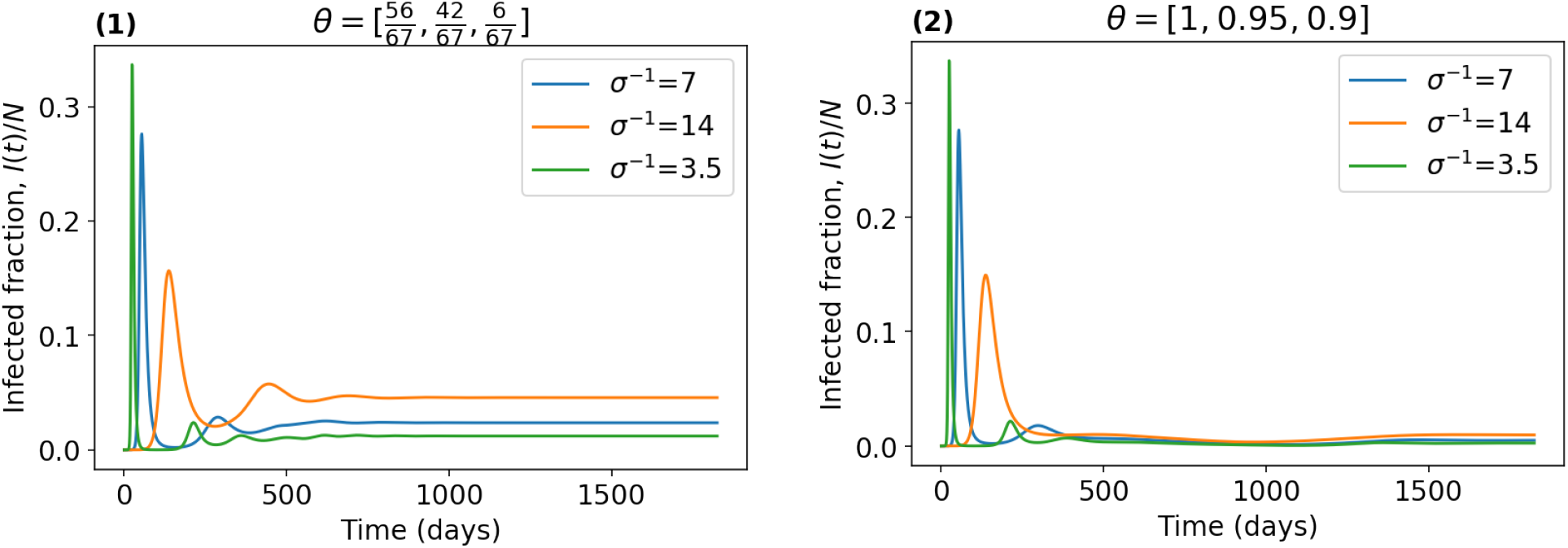
Changes in infected fraction 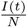 for different *σ*^−1^, and values used for other parameters are shown in Table 1.

**Figure 7:**
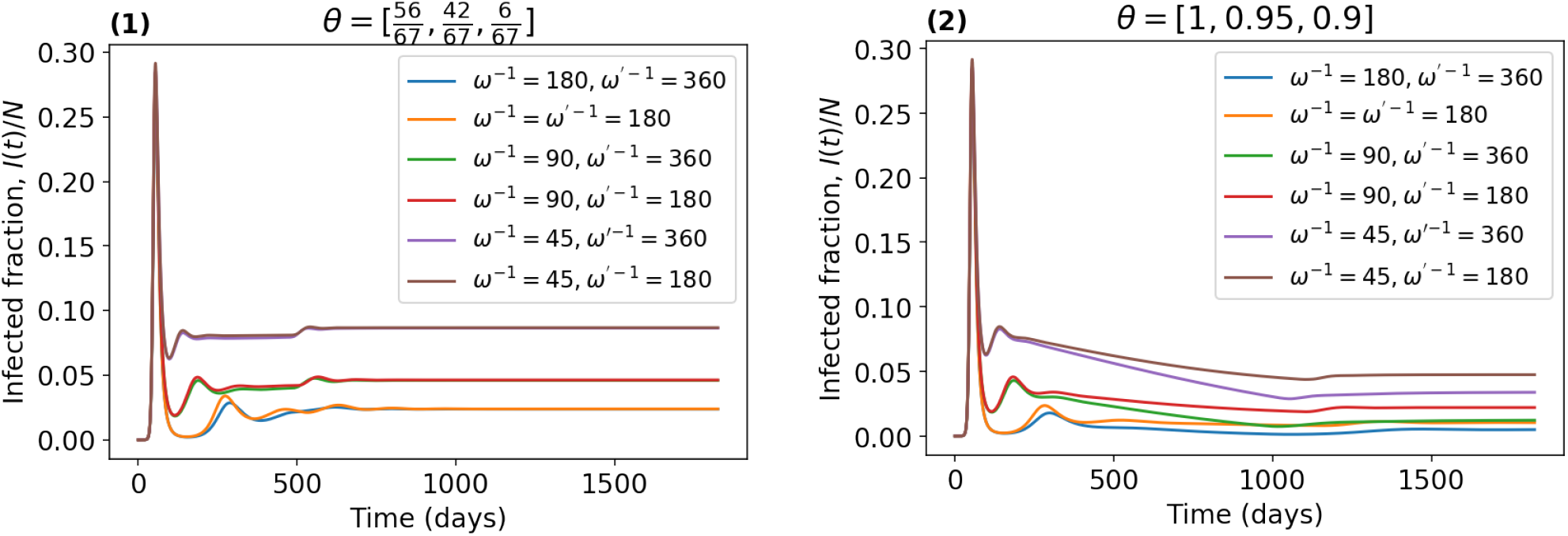
Changes in infected fraction 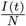 for different *ω*^−1^ and *ω*^*′*−1^, and values used for other parameters are shown in Table 1.

**Figure 8:**
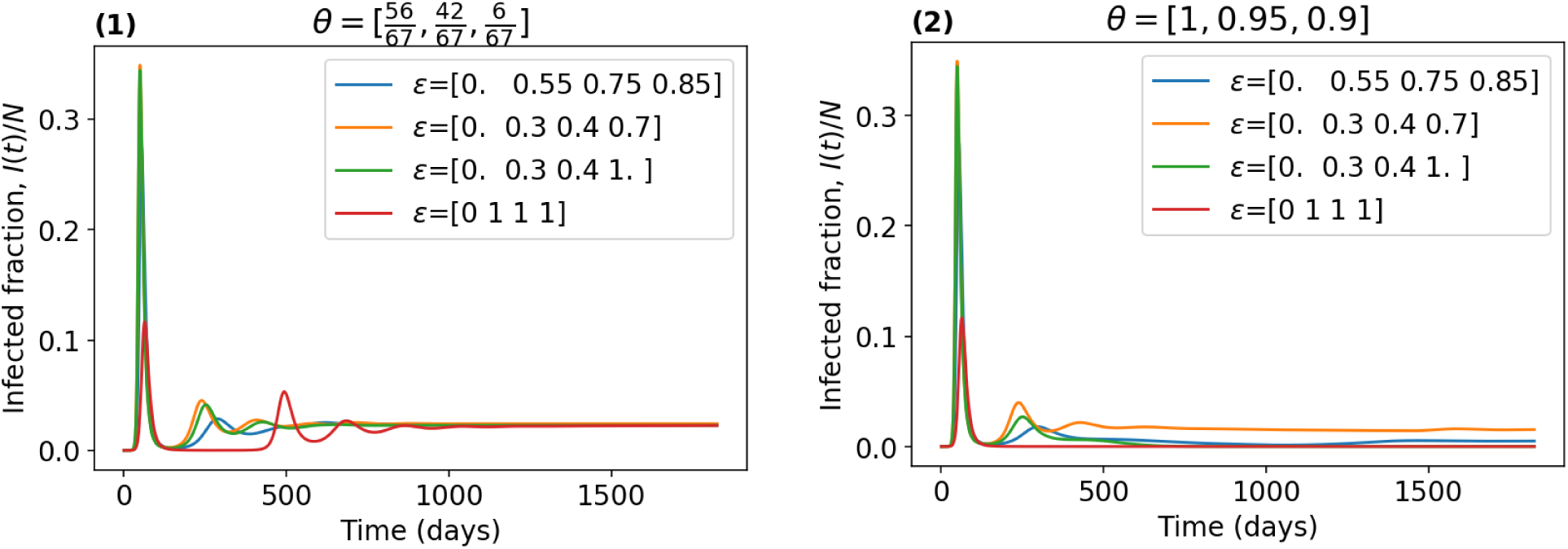
Changes in infected fraction 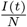 for different *ϵ*, and values used for other parameters are shown in Table 1.

**Figure 9:**
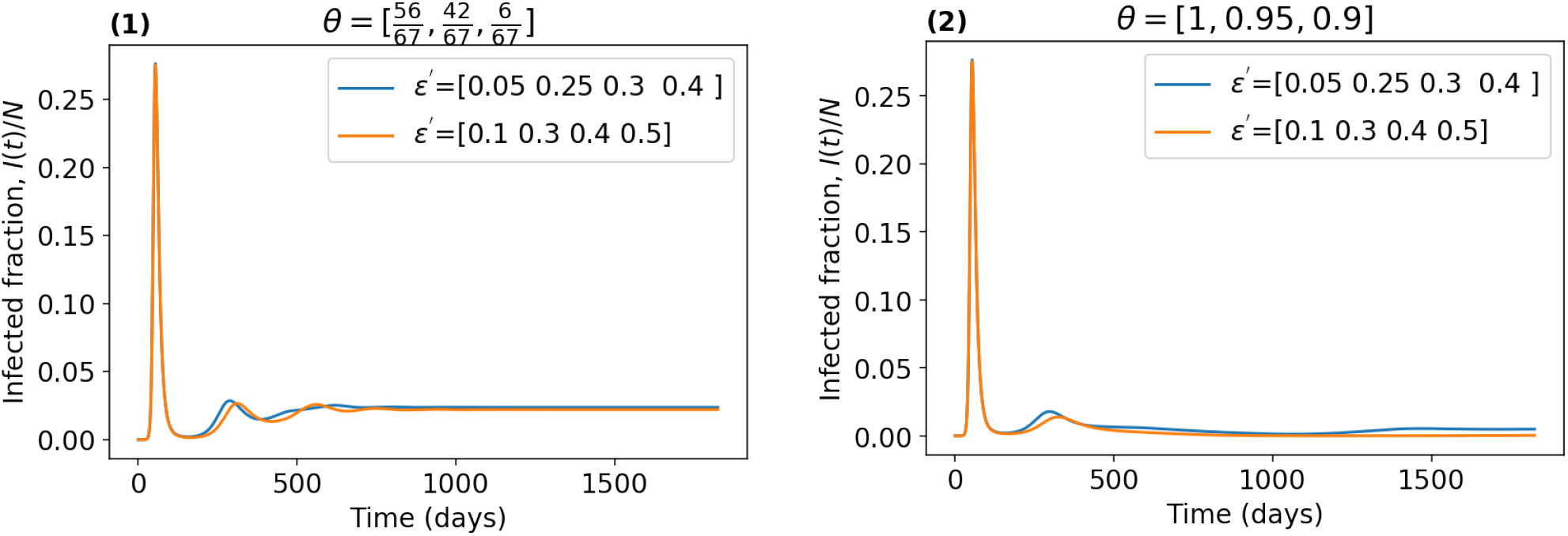
Changes in infected fraction 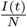 for different *ϵ*^*′*^, and values used for other parameters are shown in Table 1.

**Figure 10:**
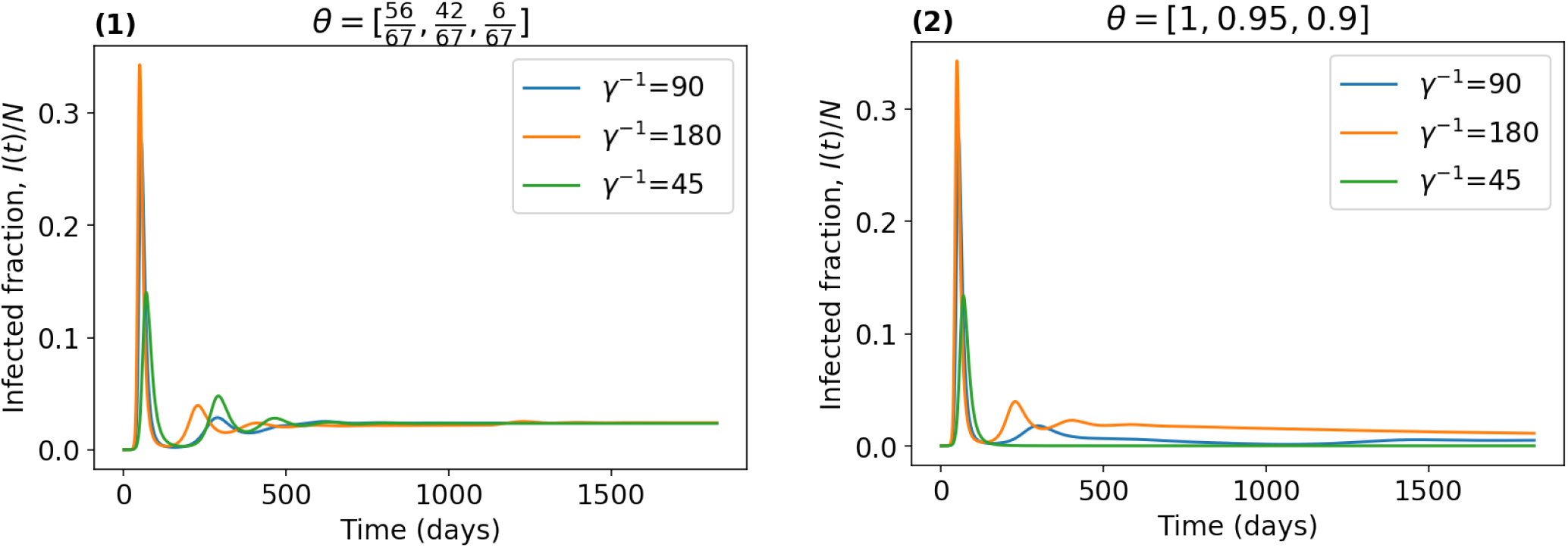
Changes in infected fraction 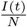 for different *γ*^−1^, and values used for other parameters are shown in Table 1.

**Figure 11:**
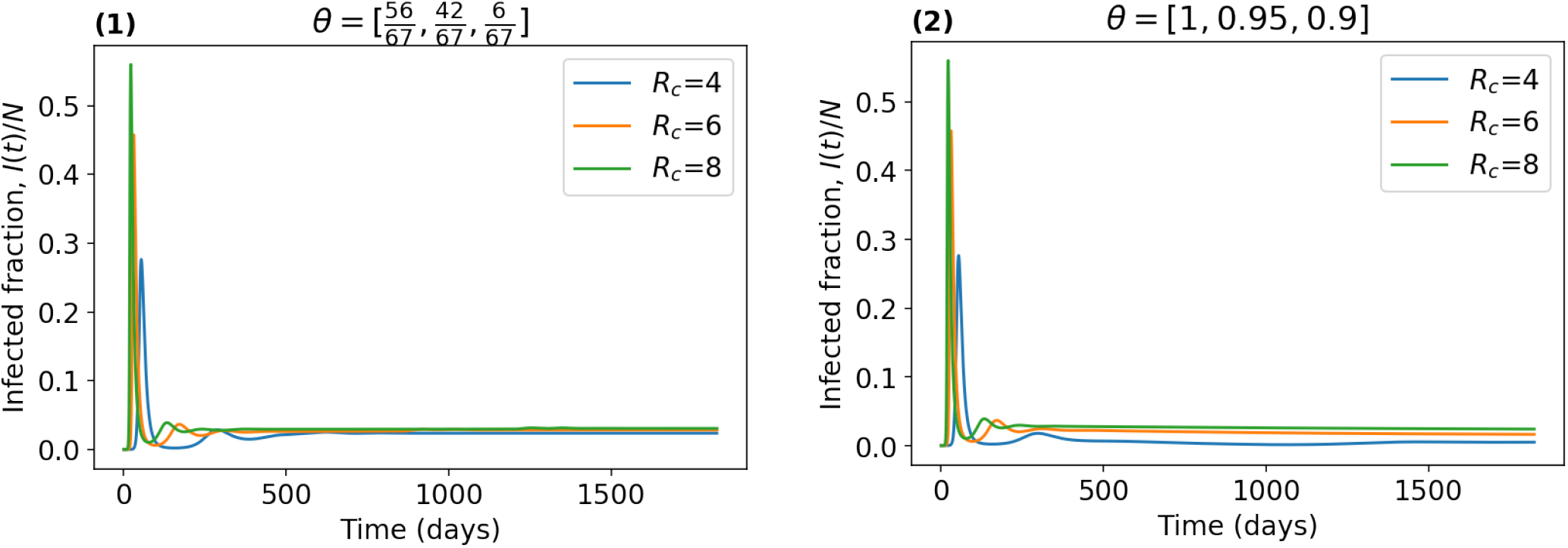
Changes in infected fraction 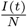 for different *R*_*c*_, and values used for other parameters are shown in Table 1.

In a), while 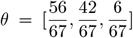, table 2 shows that both the cumulative infections over 5 years and the equilibrium incidence per year change little as the mean generation interval changes. For *θ* = [1, 0.95, 0.9], both these measures of infection are predictably smaller for corresponding values of *σ*. However, while the equilibrium incidence is quite insensitive to the value of *σ*, there is a notable reduction in cumulative infections when *σ* = 1*/*14. For both vaccination scenarios, figure 6 shows that the onset of the first wave is delayed and its peak prevalence reduces as the mean generation interval increases. Once vaccine roll out of 90% to stage is achieved, the lower equilibrium levels of infection are very apparent when compared with 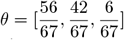.

In b) as in a), the benefit of greater vaccination coverage is again clear. Changing *ω*^*′*^ has little effect on cumulative infections when 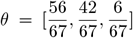, but a significant effect on all measures of infection when *θ* = [1, 0.95, 0.9]. For both vaccination scenarios, changing *ω* has a large impact on all measures of infection.

In c), we examine the effect of different vaccine efficacies, *ϵ* (before waning). It includes the cases of a perfect vaccine in stage 3, *ϵ* = (0, 1, 1, 1), (0, 0.3, 0.4, 1), for which disease free equilibrium is achieved when *θ* = [1, 0.95, 0.9]. For this scenario of high vaccination coverage, all equilibrium infection measures are highly sensitive to *ϵ*_3_. In equilibrium, it is noticeable that there is very little sensitivity to changing *ϵ* when 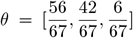. In that case, even for a perfect vaccine, endemic behaviour ensues, albeit with a very long honeymoon period between first and second waves, as figure 8 shows.

In d), we consider the effect of changing *ϵ*^*′*^. When 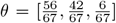, increasing *ϵ*^*′*^ by a modest amount has little effect on any of the measures of infection. But for *θ* = [1, 0.95, 0.9], table 2 shows that such a change results in a significant decline in equilibrium incidence per year and a modest one in cumulative infections over 5 years.

In e), we examine the effect of changing vaccination rates *γ*. With high vaccination coverage in stage 3, that is *θ* = [1, 0.95, 0.9], changing *γ* has a marked effect on all equilibrium measures of infection, in contrast to what happens when 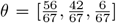.

In f), for 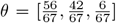, decreasing *R*_*c*_ reduces the size of the first and second waves but has only a limited effect on equilibrium incidence. The latter is in contrast to when *θ* = [1, 0.95, 0.9]. It seems that in equilibrium, increasing NPIs works best when vaccination coverage is high but has only a marginal effect when vaccination coverage is low.

In cases a) to f), figures 6 to 11 show that for each value of the parameter shown in the corresponding caption, changing from 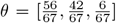 to *θ* = [1, 0.95, 0.9], has limited effect on the size and timing of the huge first waves, presumably because vaccination coverage is quite low for both these vaccination scenarios at that time.

## 6 Summary and conclusion

In this paper we develop a new vaccination model which explicitly focuses on subjects’ vaccination journeys through dose1, dose2, booster, and further boosters. The aim is to understand how infection prevalence and incidence and cumulative infections depend upon key biological parameters and control variables. We derive a single equation for calculating equilibrium prevalence in the case of endemic behaviour. We provide analytical conditions for a disease free equilibrium. We develop a numerical simulation using this compartmental vaccination model and have shown results for various parameter and control variable scenarios over a five year period. Our approach is to take a selection of parameter values constituting a ‘base case’. In section 5 we examine the effect of one at a time change in parameter values.

The key limitation of our approach is that is not age heterogeneous, and therefore, because infection fatality rates are highly age-dependent, it applies only to levels of infection, and not to Covid deaths. Secondly, many parameter values are not known with any accuracy. In future, as parameter values, including that pertaining to efficacy against transmission given infection, become known with greater certainty, and as new variants appear, the model can be run for such situations.

We examined two vaccination scenarios. The first (low aspiration) involved a population in which 84%, 63% and 9% of the population aspire to doses 1+2, booster, and repeated boosters respectively. This approximately corresponds to the vaccination coverage in the UK as of August 2022. The second scenario (high aspiration) was a hypothetical population with aspiration levels of 100%, 95%, and 90% for dose 1+2, booster, and repeated boosters, respectively.

We found that as far as equilibrium behaviour was concerned, with low vaccine aspiration, as hypothetical vaccine efficacies increased, there was no great decrease in levels of infection. The opposite was true for the high vaccine aspiration group with a disease free equilibrium actually possible for a vaccine in which repeated boosters had a hypothetical 100% efficacy. A similar effect was noticed as we changed vaccination rates. When it comes to the rate of waning immunity after vaccination, lower waning rates had little effect on equilibrium infection levels with low vaccine aspiration, but a significant effect for high vaccine aspiration. In contrast, for lower rates of waning immunity following infection, there were significant reductions in equilibrium levels of infection for both vaccination scenarios. We also found that reducing the control reproduction number (i.e increasing NPIs) had limited effect upon equilibrium infection levels with low vaccine aspiration, but a significant one with high vaccination aspiration.

The key message therefore appears to be, that for equilibrium endemic behaviour, if one wishes to achieve low levels of infection by improving any one of vaccine efficacy, waning rates, vaccination rates, and NPIs, it may only help to a significant effect if a large proportion of the population is being vaccinated on a recurrent basis.

## Data Availability

All parameter values and control variable values are illustrative and clearly stated in the article.

